# Tilted 3D visual scenes – A new therapy approach for pusher syndrome

**DOI:** 10.1101/2021.09.27.21263318

**Authors:** Sophia Nestmann, Lisa Röhrig, Björn Müller, Winfried Ilg, Hans-Otto Karnath

## Abstract

Hemiparetic stroke patients with ‘pusher syndrome’ use their non-paretic extremities to push towards their paralyzed side and actively resist external posture correction. The disorder is associated with a distorted perception of postural vertical combined with a maintained, or little deviating perception of visual upright. With the aim of reducing this mismatch, and thus reducing pushing behavior, we manipulated the orientation of visual input in a virtual reality setup. We presented healthy subjects and an acute stroke patient with severe pusher syndrome a 3D visual scene that was either upright or tilted in roll plane by 20°. By moving the sitting participants in roll plane to the left and right, we assessed the occurrence of active pushing behavior, namely the active resistance to external posture manipulation. With the 3D visual scene oriented upright, the patient with pusher syndrome showed the typical active resistance against tilts towards the ipsilesional side. He used his non-paretic arm to block the examiner’s attempt to move the body axis towards that side. With the visual scene tilted to the ipsiversive left, his pathological resistance was significantly reduced. Statistically, the tolerated body tilt angles no longer differed from those of healthy controls. We conclude that even short presentations of tilted 3D visual input can reduce pusher symptoms. The technique provides potential for a new treatment method of pusher syndrome and offers a simple, straightforward approach that can be effortlessly integrated in clinical practice.

## Introduction

An unimpaired perception of upright body orientation is crucial for balance in everyday life. Stroke patients with ‘pusher syndrome’ suffer from a specific disturbance in the perception of own body orientation (1). They perceive their bodies as ‘upright’ when actually tilted in roll plane towards the ipsilesional side (1,2) by on average 18° in the acute phase of stroke (1). Clinically, patients use their non-paretic hand (e.g., when sitting on a patient couch) or leg (when standing) to push actively towards the contralesional side until reaching postural instability and falling towards that side. External attempts to correct posture to true vertical are responded with active resistance by abducting and extending the non-paretic extremities, blocking the external movement (for review, see (3)).

Contraversive pushing occurs in around ten percent of acute stroke patients with hemiparesis (4,5) and is associated with prolonged rehabilitation times (5). A variety of therapy methods have been developed, like the use of visual feedback for upright body adjustment (6–10), machine supported gait training (11,12), lateral sitting training on a tilted plane (13,14), trunk strengthening (15), or a balance-based intervention program (16). Further approaches used stimulation techniques, like galvanic vestibular stimulation (12,17,18), transcranial direct current stimulation (17,19) or electromyographic activity-based electrical stimulation (20).

The objective of the present study was to test a new therapy approach using virtual reality (VR). The method was based on the observation that patients with pusher syndrome show a dissociation between a misaligned perception of own body orientation in space (‘subjective postural vertical’ [SPV]) (1,2) and a maintained, or little deviating perception of visual upright in relation to the body (‘subjective visual vertical’ [SVV]) (1,25). A possible explanation for the occurrence of pushing behavior has been seen in the patients’ attempt to compensate for this mismatch (3). In order to diminish the conflict between SPV and SVV percepts, we created a VR setup that allowed tilting a 3D scene in roll plane. By presenting visual input that was tilted towards the ipsilesional side of a patient with pusher syndrome, we aimed for an alignment of visual and (distorted) postural percept of verticality. Based on the reported average extend of SPV displacement in patients with pusher syndrome (1), we chose a tilt angle of 20° for the visual scene. We hypothesized that such visual manipulation might correct for postural distortions and pushing behavior in pusher syndrome.

## Methods

### Participants

The presented patient with pusher syndrome (‘P1’, letters do not refer to the patients initials) is an Italian in his late 40’s. He was found lying on the floor of his apartment and admitted to the local Centre of Neurology presenting with reduced vigilance and right-sided hemiparesis. Neuroimaging revealed a hemorrhagic stroke of the left basal ganglia combined with intraventricular hemorrhage. He underwent successful decompression surgery. A figure of the computed tomographic scans at the time of admission and 17 days post-stroke had to be removed from the manuscript due to medRxiv restrictions but can be obtained from the corresponding author on reasonable request. Motor function was assessed by the BMRC (British Medical Research Council) scale (0: no movement, 1: palpable flicker, 2: movement without gravity, 3: movement against gravity 4: movement against resistance, 5: normal movement). His right arm was plegic (0/5), his right leg severely paretic (proximal: 2/5, distal: 1/5). He presented with severe pusher syndrome, as assessed by the ‘Scale for Contraversive Pushing (SCP)’ (1,26,27) 27 days after admission and at the time of experimental testing 29 days after admission. SCP results were unchanged between the two time points of testing with scores of 2 (‘Spontaneous body posture’) and 2 (‘Use of the non-paretic extremities [abduction & extension]’). ‘Resistance to passive correction of tilt posture’ could only be tested during sitting and showed a value of 1 at both time points. Neurological examination revealed no clinical signs of visual extinction, spatial neglect, or visual field defects; P1 suffered from aphasia and understood only rudimentary instructions in Italian. Formal neuropsychological testing was not possible.

As a control sample, we included 13 older subjects (mean age: 67.5 ± 6.6 [SD] years; 4 females) in order to provide a baseline of healthy individuals’ behavior during our experimental manipulation. The experiment was conducted in accordance with the Declaration of Helsinki and was approved by the ethics committee of the medical faculty, University of Tübingen. All participants gave written informed consent before participation.

**Fig. 1.** Computed tomography scans of P1’s brain showing. **A)** a hyperdense hemorrhagic stroke of the left basal ganglia and intraventricular hemorrhage with midline shift before decompression surgery at admission, and **B)** the lesion 17 days after surgery. The figure had to be removed due to medRxiv policy but can be obtained from the corresponding author on request.

### Stimuli and Procedure

Subjects were seated on a patient couch with their feet hanging freely. Via a head-mounted-display (HTC Vive, HTC Corporation, Taoyuan City, Taiwan), they were presented with the scene of a beach (adapted from (28); cf. Fig. 2A). Note that in contrast to the monocular view in Figure 2A, the presented scene was three-dimensional with the participants seated on the footbridge. They were instructed to naturally view the presented scene and not to close their eyes during the experiment.

**Fig. 2.**
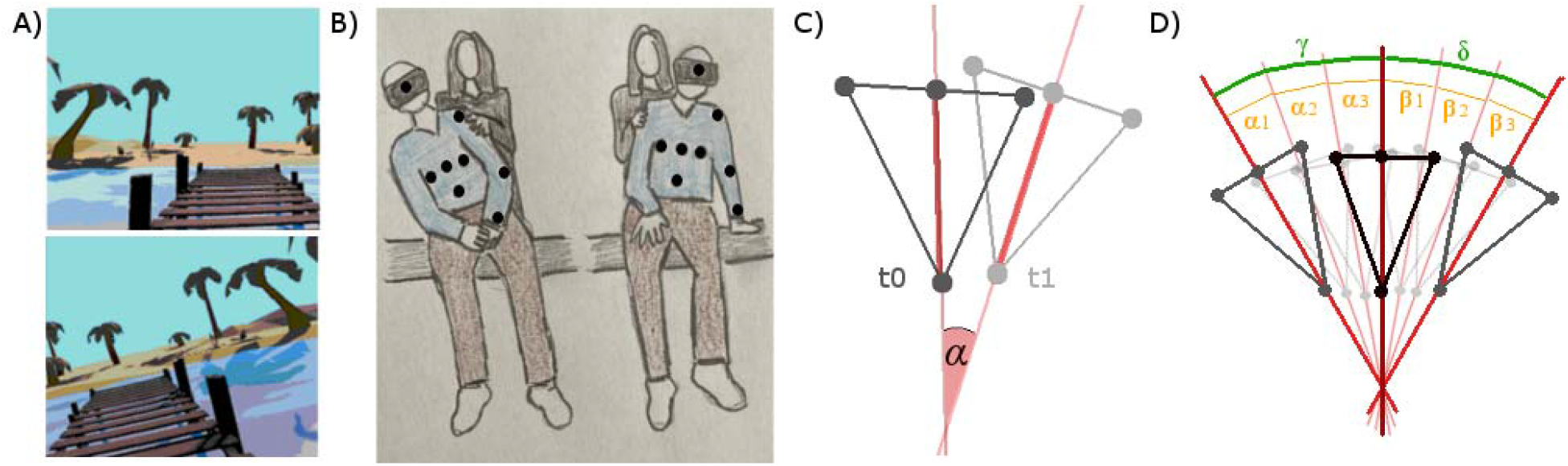
Experimental setup. (**A**) The virtual environment presented via a head-mounted-display. The scene was presented either aligned with the true vertical or tilted in roll plane. Note that in contrast to the monocular view in this figure, in VR the presented scene was three-dimensional, and participants were located on the footbridge. (**B**) The experimenter stood behind the participants and moved them at their shoulders to the left and to the right [due to the medRxiv policy we may only display a sketch of the setup, the original picture can be obtained from the corresponding author on request]. Spherical reflective markers attached to the subjects’ trunk as well as to shoulder, elbow, and wrist allowed motion capturing. Their rough location is indicated by black dots. In the right panel, the active resistance behavior of pusher patient P1 (using left arm and hand) that occurred for tilt movements towards his left is illustrated. **(C)** Schematic illustration of the determination of body tilt from the position of VICON body markers in two consecutive trials. Dark colors represent the markers’ positions in an initial trial t0; light colors in the consecutive time point t1. The three dots in a row represent the markers applied at a participant’s chest [left, middle, right]. The single dot below represents the marker applied approximately at the belly button. As indicator for the body’s longitudinal axis, we determined the vector between the mid-chest marker and the marker located at the belly button (red line). The change in body orientation was calculated as the angle α between this vector at two consecutive time points t0 and t1. **(D)** Resulting angles from each time point of each trial were cumulated in order to receive the overall amount of body tilt to the left and right. In the displayed example, the overall tilt γ would be calculated as γ = α1 + α2 + α3 and the angle δ as δ = ß1 + ß2 + ß3.

In P1, the experiment consisted of three consecutive conditions: In ‘condition 1’, the scene was aligned with the true vertical, in ‘condition 2’, the scene was tilted by 20° to the ipsiversive left (cf. Fig. 2A), and in ‘condition 3’, the scene was again aligned with the true vertical. The center of rotation was located 0.5m below the coordinates of the headset, which in a person of average height is slightly above the belly button. In controls, conditions 1 and 3 were performed identically. In condition 2, the direction of scene tilt was balanced across subjects in order to avoid any bias potentially associated with tilt side. This was done by presenting a scene tilted by 20° to the left in half of the sample and 20° to the right in the other half. Each condition consisted of ten trials. Due to P1’s severe medical condition, we reduced trials for him by only recording five trials in condition 1 and seven trials in condition 3.

P1 suffered from severe pusher syndrome and was not able to sit unassisted without pushing himself to the side and falling over. We thus assessed the effect of experimental intervention on the active resistance towards external attempts of posture manipulation towards the ipsilesional side, which is the typical behavior of patients with pusher syndrome. In each trial, the experimenter stood behind the participants and moved them in roll plane to the left and to the right, securely holding the subjects at their shoulders (Fig. 2B). Participants were always aware that the experimenter would prevent falling and (in line with the instructions from the SCP) they were asked to allow the experimentally induced movements. Start and end positions were always at the upright body orientation. At the beginning of each trial, the hands were positioned on the lap, but participants were intentionally not encouraged to keep their hands on the lap at all costs, as this would have contradicted the experimentally intended possibility of using the hands to resist postural manipulation. Tilt movements were terminated either by the subject by actively resisting against more extreme body tilts (see right panel of Figure 2B for an example) or by the experimenter who did not exceed lateral tilt angles of 30° to 40°. In P1, the initial movement in all three conditions was towards the contraversive right, in healthy subjects always towards the direction of scene tilt chosen in condition 2. The duration of each trial was 9.0 ± 2.8 seconds in P1 and 5.4 ± 0.7 in controls.

Participants’ longitudinal body axis was continuously tracked with a VICON MX-3-Series motion capturing system (Vicon Motion Systems Ltd, Yarnton, United Kingdom) using five infrared cameras. We attached one spherical reflective marker to the headset, three horizontally at the chest and one at the above location of the belly button (see Fig. 2B). Further markers were applied to both shoulders, elbows, and wrists to be able to capture the typical active resistance of patients with pusher syndrome to posture manipulation towards the ipsilesional side (7).

### Data analysis

The VICON Nexus Software (http://www.vicon.com/products/software/nexus) was used for recording and pre-processing. Motion was captured at a sample rate of 250Hz. We applied the core processing routine and filled trajectory gaps using a Woltering filter. Figure 2C schematically illustrates the determination of participants’ body orientation from the position of the VICON markers. As indicator of body orientation, we determined the vector between the mid-chest marker and the marker located at the belly button. Mathematically, the change in body orientation was calculated as the angle α between this vector at time point t0 and t1 and all subsequent time points. The initial starting position was determined as participant’s average upright orientation for trials of condition 1. For each trial, data were smoothed by excluding time points, where the calculated angle differed more than two standard deviations from the mean. Resulting angles were cumulated, resulting in an overall amount of body tilt for movements to the left and right in each trial. A schematic illustration is shown in Figure 2D, where the overall tilt γ is calculated as γ = α1 + α2 + α3 and the angle δ is calculated as δ = ß1 + ß2 + ß3.

In healthy controls, we excluded single trials of participants as outliers if the calculated tilt angle differed more than two standard deviations from the average of all trials in all participants and conditions (n = 13). One further trial had to be excluded because of technical constraints. The remaining dataset consisted of 376 trials. In order to avoid removing information specific for pusher syndrome, we did not exclude any trials from P1’s data. His data set consisted of tilt angles from 22 trials, which were averaged per condition and tilt side.

For better comparability of P1’s and control subjects’ data, we coded movements towards the direction of scene tilt as ‘contralesional’ and movements away from the direction of scene tilt as ‘ipsilesional’. To check, whether the experimentally induced tilt in the group of healthy controls differed significantly between the conditions, we calculated a mixed ANOVA with the *aov()* function provided by R’s *stats* package (29). We included the between-subjects factors *Visual Scene Orientation* (‘Baseline Upright’, ‘Tilted’, ‘Post Upright’) and *Tilt Side* (‘Ipsilesional’, ‘Contralesional’), as well as their interaction. Repeated measurements were accounted for by including the within-subjects factor *Participant*. Data were centered at the baseline mean by subtracting the ipsilesional mean in condition 1 from all ipsilesional trials and the contralesional mean from contralesional trials accordingly. To assess patient P1’s behavior in relation to the control group, we used the Crawford-Garthwaite Bayesian test for single-case analysis (30) as implemented in R’s *psycho* package (31).

## Results

In the group of healthy controls, none of the subjects resisted experimental tilt movements in any trial of the three conditions. Tilt angles thus corresponded with the manipulation applied by the experimenter. As revealed by a mixed ANOVA, we observed no statistically significant effect of tilt side (*F* = 0.37, *p* = 0.545) or condition (*F* = 1.54, *p* = 0.223) on the amount of experimentally applied body tilt. P1 did not show resistance for tilts towards his contralesional side. However, as expected, he resisted body tilting towards his ipsilesional side by moving his non-paretic hand from the lap to the patient couch, blocking the movement by abducting and extending the left arm. This forceful resistance was observed in each trial of ipsiversive tilt and was documented through the VICON markers positioned at the patient’s shoulder, elbow, and wrist.

As the experimental intervention aimed at reducing resistance in P1, we assessed a potential normalization of his behavior in the second and third condition by comparing his data to the baseline of healthy controls. In baseline condition 1, controls were tilted on average 33.07° to the right and 32.44° to the left; the amount of tilt to the left and right did not differ significantly (paired t-test: *t* = 0.41, *p* = 0.683). For further analyses, we thus calculated an overall average tilt angle (32.75°, *SD* = 11.08), which the patient’s data was compared to by using the Crawford-Garthwaite Bayesian test for single-case analysis (30). Data are illustrated in Figure 3.

**Fig. 3.**
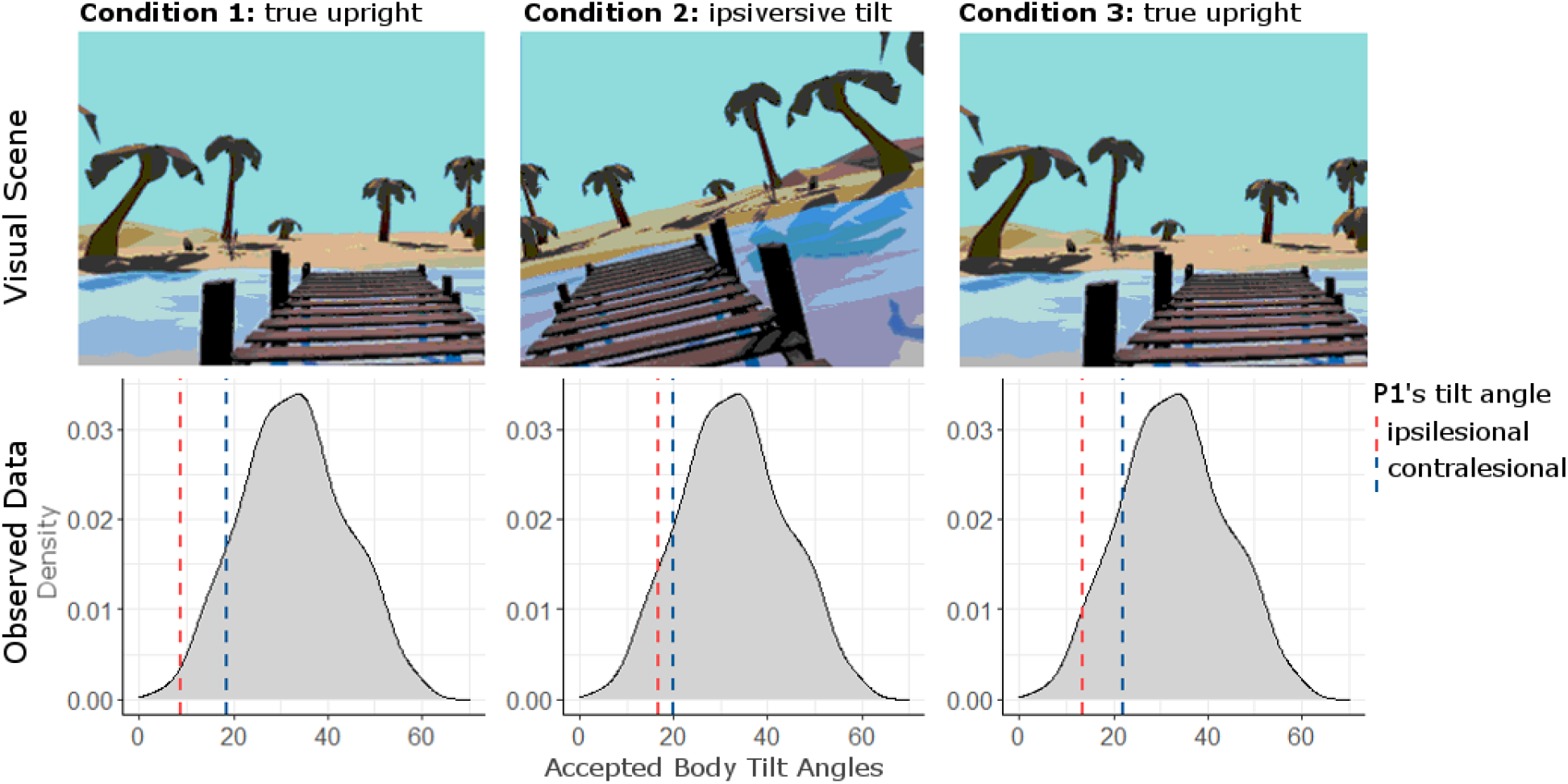
Visual scene presented via the head-mounted-display and corresponding results. Density plots of healthy controls’ baseline data with body tilts of the patient P1 in the three experimental conditions. Tilt angles are displayed on the x-axis. Patient P1’s averaged contralesional body tilt is represented by blue dashed vertical lines, his averaged body tilts towards the ipsilesional side by red dashed lines. P1 blocked movements to the ipsilesional side in each trial of each condition.

In condition 1, patient P1’s body tilt towards the contralesional side did not differ statistically significant from healthy controls’ data (P1: 18.42°, *SD* = 4.23°, *Z* = −1.29, *p* = 0.119). In contrast, he allowed significantly smaller tilts towards his ipsilesional side (P1: 8.77°, *SD* = 3.00°; *Z* = −2.17, *p* = 0.030). With the VR scene tilted by 20° towards his ipsilesional side (condition 2), we observed a reduction of P1’s resistance towards the experimentally applied ipsiversive tilt. P1 now allowed tilts of on average 16.90° (*SD* = 3.14°) to his ipsilesional side and 19.91° (*SD* = 1.85°) to his contralesional side. Statistically, P1’s ipsiversive body tilt did no longer differ significantly from the control subjects’ baseline in condition 1 (*Z* = −1.43, *p* = 0.095). In condition 3, in which the VR-scene was again aligned with the true vertical, we observed an aftereffect of our intervention: P1 allowed an average ipsiversive tilt of 13.60° (*SD* = 5.46°) which was not significantly different from the controls’ baseline (*Z* = −1.73, *p* = 0.060). Towards his contralesional side, he was moved by on average 22.11° (*SD* = 4.15°).

## Discussion

The presented experiment aimed at reducing the mismatch between visual and postural percepts of upright in pusher syndrome that had been observed in previous studies (1,2,25). We observed that tilting a 3D visual scene towards a patient’s ipsilesional side by 20° reduced his pathological resistance against external posture manipulations. Under this experimental condition, body tilt angles towards the ipsilesional side no longer differed statistically significant between the patient with severe pusher syndrome and healthy controls.

The brain is believed to generate a percept of upright as the weighted average of available information about verticality (33–36). In the case of experimentally induced misalignment of visual and postural verticality information, this integration results in a bias of body upright orientation in the direction of visual scene tilt, especially pronounced in older subjects (37). This observation is in line with the previously reported prominent importance of visual information for the perception of upright (38,39). It also corresponds to the alterations in visual and postural percepts of verticality, caused by a rotation of the visual surroundings (40,41). Nonetheless, integrative processes seem to be based on only a subset of reliable information, when information about verticality differs too much between modalities (36).

Based on these insights, the misalignment between the SPV and SVV of a patient with pusher syndrome should result in an overall body orientation aligned with an intermediate orientation between postural and visuo-vestibular percept or □ if the model of partial integration holds □ with either the SPV or SVV. However, patients with pusher syndrome align their bodies with neither of them but push towards the contralesional side (1). While at the first glance this seems contradictive, a closer look reveals that pusher syndrome differs significantly from experimental manipulations induced in healthy participants. Experiments with healthy controls only allow a manipulation of visual input, while in patients with pusher syndrome the SPV is distorted. Based on these considerations, adaptations of body posture towards the true upright orientation are conceivable if visual and postural percepts become re-aligned as in our experiment. This would reduce the diversity of available verticality information (3,24) and, in consequence, should decrease pushing behavior. Our very promising observations in patient P1 support this hypothesis and offer a simple, straightforward therapy approach.

At a first glance, our present findings might seem different from observations in a recent study by Mansfield et al. (32). Among other stroke patients and healthy controls, the authors investigated one patient with active pushing behavior for his ability to remain seated upright on a motion platform, which was oriented either upright or tilted to the ipsi- or contralesional side in roll plane by 18°, with or without a (consistently) tilted surrounding visual scene. Interestingly, participants with stroke - including the one patient with active pushing behavior - did not appear to adjust their posture in response to visual scene tilt during upright motion platform to a greater extent than control participants. However, participants were seated on a plinth without anything near their hands or feet that they could have touched and hence used to actively push to the side. In other words, their setting did not allow the participating patient with pusher syndrome to show the pathological behavior of active pushing to the side, namely the use of the non-paretic arm or leg to cause the pathological lateral tilt of the body. The latter is only observed if patients with pusher syndrome can make contact to the ground, i.e. to the surface of a bed, patient couch, or to the floor (6,7). The possibility of making contact to the ground is also the decisive feature to provoke and observe the typical blocking behavior of patients with pusher syndrome: any external manipulation aiming at shifting the body weight towards the non-paretic side elicits active resistance by the patients, who increase the force of the extended non-paretic extremity, blocking any movement (6,7). In contrast to the setup used by Mansfield et al. (32), our present experiment was especially designed to allow and provoke these features of active pushing behavior: participants were free to use hands and arms to make contact to the ground and block external posture manipulations.

It will be interesting for future studies to investigate long-term effects of immediate VR-driven perceptional alterations (42). The same applies to possible side effects caused by a longer exposure to tilted 3D visual scenes, like motion sickness, that could occur during or after treatment. If necessary, parameters will need to be adjusted for a viable therapeutic approach. Two further aspects related to future studies should be considered. First, P1 suffered from severe pusher syndrome and was not able to sit unsupported. We were therefore only able to investigate the effect of intervention on active resistance against posture manipulation. This important aspect of pusher syndrome plays an essential role during the rehabilitation process of pusher syndrome and thus receives justified attention. Nevertheless, the presented treatment method should also be applied to less impaired patients in future research. We expect that in such patients our treatment method allows unsupported sitting by reducing the urge of compensating the experienced mismatch in verticality perception. Second, because of P1’s aphasia, it was not possible to instruct him sufficiently, which would have allowed us to determine the exact extent of his individual SPV perception on a lateral tilt chair. With this value, it would have been possible to individually adjust the tilt angle of the visual scene. Instead, we chose an ipsiversive tilt of the visual scene of 20°, which corresponded roughly to the previously measured average misalignment of SPV in a group of acute stroke patients with pusher syndrome (1). As originally intended, we suggest that future studies using our approach should choose a degree of visual scene tilt that corresponds to the degree of individual SPV distortion.

To conclude, in the present experiment we observed a reduction of pushing behavior when a patient was presented with a 3D VR scene that was tilted to the ipsilesional side in roll plane. Our single case study suggests that VR might become a promising tool in the treatment of pusher syndrome to reduce rehabilitation time and improve functional mobility.

## Data Availability

Data analyzed during the current study are not publicly available due to the data protection agreement of the Centre of Neurology at Tübingen University, as approved by the local ethics committee and signed by the participants. They are available on reasonable request to the corresponding author following completion of a formal data sharing agreement, after obtaining formal informed consent of each participant for the use of his/her data on this purpose, and approval by the local ethics committee.

## List of abbreviations

VR: Virtual Reality
SPV: Subjective Postural Vertical
SVV: Subjective Visual Vertical

## Acknowledgements

This work was supported by the Deutsche Forschungsgemeinschaft (KA 1258/23-1). We thank patient P1 for his willingness to participate in our experiment. We also thank Tobias Schumacher, Marion Himmelbach, and Franziska Oesterle for their support during patient recruitment, as well as Endi Epifanio and Felix Müller for physiotherapeutic assistance during the experiment and for translating when communicating with the patient.

## References

1. Karnath, H. O., Ferber, S., & Dichgans, J. (2000). The origin of contraversive pushing: Evidence for a second graviceptive system in humans. Neurology, 55(9), 1298–1304. https://doi.org/10.1212/wnl.55.9.1298

2. Bergmann J, Krewer C, Selge C, Müller F, Jahn K. The Subjective Postural Vertical Determined in Patients with Pusher Behavior During Standing. Topics in Stroke Rehabilitation. 2016 Apr 2;23(3):184–90.

3. Karnath H-O. Pusher Syndrome – a frequent but little-known disturbance of body orientation perception. Journal of Neurology. 2007 Apr;254(4):415–24.

4. Abe H, Kondo T, Oouchida Y, Suzukamo Y, Fujiwara S, Izumi S-I. Prevalence and length of recovery of pusher syndrome based on cerebral hemispheric lesion side in patients with acute stroke. Stroke. 2012 Jun;43(6):1654–6.

5. Pedersen PM, Wandel A, Jørgensen HS, Nakayama H, Raaschou HO, Olsen TS. Ipsilateral pushing in stroke: incidence, relation to neuropsychological symptoms, and impact on rehabilitation. The Copenhagen Stroke Study. Arch Phys Med Rehabil. 1996 Jan;77(1):25–8.

6. Broetz D, Karnath H-O. New aspects for the physiotherapy of pushing behaviour. NeuroRehabilitation. 2005;20(2):133–8.

7. Karnath H-O, Broetz D. Understanding and Treating “Pusher Syndrome.” Physical Therapy. 2003 Dec 1;83(12):1119–25.

8. Luque-Moreno C, Jiménez-Blanco A, Cano-Bravo F, Paniagua-Monrobel M, Zambrano-García E, Moral-Munoz JA. Effectiveness of visual feedback and postural balance treatment of post-stroke pusher syndrome. A systematic review. Revista Científica de la Sociedad de Enfermería Neurológica (English ed) [Internet]. 2020 Apr 22 [cited 2021 Mar 26]; Available from: https://www.sciencedirect.com/science/article/pii/S2530299X20300030

9. Yang Y-R, Chen Y-H, Chang H-C, Chan R-C, Wei S-H, Wang R-Y. Effects of interactive visual feedback training on post-stroke pusher syndrome: a pilot randomized controlled study. Clin Rehabil. 2015 Oct 1;29(10):987–93.

11. Bergmann J, Krewer C, Jahn K, Müller F. Robot-assisted gait training to reduce pusher behavior: A randomized controlled trial. Neurology. 2018 Oct 2;91(14):e1319–27.

13. Fukata K, Amimoto K, Inoue M, Sekine D, Inoue M, Fujino Y, et al. Effects of diagonally aligned sitting training with a tilted surface on sitting balance for low sitting performance in the early phase after stroke: a randomised controlled trial. Disability and Rehabilitation. 2019 Nov 12;0(0):1–9.

14. Fukata K, Amimoto K, Inoue M, Shida K, Kurosawa S, Inoue M, et al. Effects of performing a lateral-reaching exercise while seated on a tilted surface for severe post-stroke pusher behavior: A case series. Topics in Stroke Rehabilitation. 2020 Dec 20;1–8.

15. Saeys W, Truijen S. The effect of trunk exercises on the perception of verticality after stroke: A pilot study. Neurol Rehabil. 2019;25:S37–41.

16. Pardo V, Galen S. Treatment interventions for pusher syndrome: A case series. NeuroRehabilitation. 2019 Jan 1;44(1):131–40.

17. Babyar S, Santos T, Will-Lemos T, Mazin S, Edwards D, Reding M. Sinusoidal Transcranial Direct Current Versus Galvanic Vestibular Stimulation for Treatment of Lateropulsion Poststroke. Journal of Stroke and Cerebrovascular Diseases. 2018 Dec 1;27(12):3621–5.

18. Nakamura J, Kita Y, Yuda T, Ikuno K, Okada Y, Shomoto K. Effects of galvanic vestibular stimulation combined with physical therapy on pusher behavior in stroke patients: A case series. NeuroRehabilitation. 2014 Jan 1;35(1):31–7.

19. Yamaguchi T, Satow T, Komuro T, Mima T. Transcranial Direct Current Stimulation Improves Pusher Phenomenon. Case Rep Neurol. 2019 Feb 28;11(1):61–5.

20. Fujino Y, Takahashi H, Fukata K, Inoue M, Shida K, Matsuda T, et al. Electromyography-guided electrical stimulation therapy for patients with pusher behavior: A case series. NeuroRehabilitation. 2019 Jan 1;45(4):537–45.

21. Bisdorff AR, Wolsley CJ, Anastasopoulos D, Bronstein AM, Gresty MA. The perception of body verticality (subjective postural vertical) in peripheral and central vestibular disorders. Brain. 1996 Oct;119 (Pt 5):1523–34.

22. Funabashi M, Santos-Pontelli TEG, Colafêmina JF, Pavan TZ, Carneiro AAO, Takayanagui OM. A new method to analyze the subjective visual vertical in patients with bilateral vestibular dysfunction. Clinics (Sao Paulo). 2012 Oct;67(10):1127–31.

23. Kobayashi H, Hayashi Y, Higashino K, Saito A, Kunihiro T, Kanzaki J, et al. Dynamic and static subjective visual vertical with aging. Auris Nasus Larynx. 2002 Oct;29(4):325–8.

24. Perennou DA, Mazibrada G, Chauvineau V, Greenwood R, Rothwell J, Gresty MA, et al. Lateropulsion, pushing and verticality perception in hemisphere stroke: a causal relationship? Brain. 2008 Aug 21;131(9):2401–13.

25. Johannsen L, Fruhmann Berger M, Karnath H-O. Subjective visual vertical (SVV) determined in a representative sample of 15 patients with pusher syndrome. Journal of Neurology. 2006 Oct;253(10):1367–9.

26. Baccini M, Paci M, Nannetti L, Biricolti C, Rinaldi LA. Scale for contraversive pushing: cutoff scores for diagnosing “pusher behavior” and construct validity. Phys Ther. 2008 Aug;88(8):947–55.

27. Karnath H-O, Brötz D. Instructions for the Clinical Scale for Contraversive Pushing (SCP). Neurorehabil Neural Repair. 2007 Aug;21(4):370–1; author reply 371.

28. Moritz J-M. Ganzkörper-Repräsentation in der Virtuellen Realität für Anwendungen in der Neurorehabilitation [Unpublished master’s thesis]. Eberhard Karls Universität Tübingen, Tübingen; 2017.

29. R Development Core Team. R: A Language and Environment for Statistical Computing [Internet]. Vienna, Austria: R Foundation for Statistical Computing; 2018. Available from: http://www.R-project.org

30. Crawford JR, Garthwaite PH. Comparison of a single case to a control or normative sample in neuropsychology: Development of a Bayesian approach. Cognitive Neuropsychology. 2007 Jun;24(4):343–72.

31. Makowski D. The psycho Package: an Efficient and Publishing-Oriented Workflow for Psychological Science. The Journal of Open Source Software. 2018 Feb 5;3(22):470.

32. Mansfield A, Taati B, Danells CJ, Fraser LE, Harris LR, Campos JL. Postural orientation with conflicting visual and graviceptive cues to ‘upright’ among individuals with and without a history of post-stroke ‘pushing.’ Neurol Rehabil. 2019 May;25:S26–32.

33. MacNeilage PR, Banks MS, Berger DR, Bülthoff HH. A Bayesian model of the disambiguation of gravitoinertial force by visual cues. Exp Brain Res. 2007 May;179(2):263–90.

34. Alberts BBGT, de Brouwer AJ, Selen LPJ, Medendorp WP. A Bayesian Account of Visual-Vestibular Interactions in the Rod-and-Frame Task. eNeuro. 2016 Oct;3(5).

35. Mittelstaedt H. A new solution to the problem of the subjective vertical. Naturwissenschaften. 1983 Jun;70(6):272–81.

36. de Winkel KN, Katliar M, Diers D, Bülthoff HH. Causal Inference in the Perception of Verticality. Scientific Reports. 2018 Apr 3;8(1):5483.

37. Nestmann S, Karnath H-O, Bülthoff HH, Nikolas de Winkel K. Changes in the perception of upright body orientation with age. Baurès R, editor. PLoS ONE. 2020 May 29;15(5):e0233160.

38. Asch SE, Witkin HA. Studies in space orientation; perception of the upright with displaced visual fields. J Exp Psychol. 1948 Jun;38(3):325–37.

39. Howard IP, Hu G. Visually induced reorientation illusions. Perception. 2001;30(5):583–600.

40. Dichgans J, Held R, Young LR, Brandt T. Moving visual scenes influence the apparent direction of gravity. Science. 1972 Dec 15;178(4066):1217–9.

41. Witkin HA. Perception of body position and of the position of the visual field. Psychological Monographs: General and Applied. 1949;63(7):i–46.

42. Wright WG. Using virtual reality to augment perception, enhance sensorimotor adaptation, and change our minds. Front Syst Neurosci. 2014;8:56.

